# Barriers and facilitators for implementing a dementia caregiver support strategy among Latinos in primary care

**DOI:** 10.1101/2025.06.25.25330304

**Authors:** Jaime Perales-Puchalt, Emily Morrow, Maria Val Roche-Dean, Ton Miras, Angela Scott, Mariana Ramirez-Mantilla, Ladson Hinton, Dolores Gallagher-Thompson, Kristi Williams, Jeffrey M Burns, Edward Ellerbeck

## Abstract

**Introduction:** Dementia has a devastating impact on Latino family caregivers, but access to caregiver support is limited. Community health workers could be leveraged to provide culturally tailored education and resources. We aimed to identify barriers and facilitators to implementation of an evidence-based and culturally-tailored caregiver support program consisting of handing an educational comic-like booklet (i.e. ¡Unidos Podemos!) supplemented with small group discussions and follow-up calls by community health workers to Latinos in primary care.

**Methods:** We conducted qualitative interviews with 20 community health workers, primary care providers, and other clinic staff recruited nationally via purposive sampling. We organized the transcripts for qualitative review to identify themes, using deductive analysis methods.

**Results:** Within the five a priori domains derived from the Consolidated Framework for Implementation Research, 21 themes were identified. These themes described a variety of barriers and facilitators to implementation of the intervention. Staff and clinicians valued the program highly and delivering it via the comic-like booklet was seen as simple. To make the program more implementable, recommendations were made to address several components that supplement and enhance use of the booklet. This included the identifying reimbursement options to schedule and hold group discussions, and incorporating staff training, decision support tools, supervision, and feedback mechanisms.

**Conclusion:** ¡Unidos Podemos! provides a unique opportunity for primary care clinics to care for their Latino caregivers of people with dementia. Findings can inform the development, refinement and testing of an implementation strategy for community health workers to deliver ¡Unidos Podemos! in primary care.

## Introduction

Family caregiving for individuals with dementia (IWDs) has a serious emotional, physical and financial toll.^1–8^ The impact of Alzheimer’s disease and related dementias (ADRD) among Latinos is high. Latinos are the largest and fastest growing elder group and are 1.5 times more likely to develop ADRD than non-Latino Whites. ^2,9–12^ Correspondingly, the number of Latino individuals with ADRD is projected to increase from 379,000 in 2012 to 3.5 million by 2060.^9^ Additionally, the prevalence and severity of neuropsychiatric symptoms among Latinos with ADRD is higher than among non-Latino Whites.^13–16^ In fact, Latinos provide more intense and longer caregiving, experience disproportionate levels of caregiver stress and depression, and have poor access to caregiver community resources. ^4,8,13,17–28^ Latinos are highly likely to become family caregivers and especially vulnerable to its negative consequences.^23,29,30^

Caregiver support programs can reduce the negative impact of ADRD on Latino caregivers. Nearly 300 caregiver programs have been tested in randomized designs^1,31–33^ to provide caregiving knowledge and skills, link caregivers to available community resources, and provide coping mechanisms for stress.^31^ Many programs are effective in reducing caregiver stress, depression and other negative outcomes.^1,31–33^ In fact, a recent systematic review identified 23 studies of non-pharmacologic programs for Latino caregivers of people with ADRD. One caregiver support program that holds promise in the Latino community is ¡Unidos Podemos!, a culturally tailored program that uses Social Learning and Health Behavior Model principles to support caregivers of IWDs.^34,35^ the program is based on a fotonovela, which is a pictorial soap opera-like storytelling booklet popular among Latinos. The ¡Unidos Podemos! fotonovela is short, as it has only 20 pages in English and Spanish. The fotonovela was developed by a team of professionals with experience in Latino dementia caregiving, to provide information about the caregiving process, including common challenges and ideas for action to reduce caregiver stress. Scripts were developed by a multidisciplinary team, forward and back translated, and then each page of the fotonovela was filmed. This work was led by an individual who had prior experience creating fotonovelas for the US Centers for Disease Control where they are widely used to transmit health information to lower literacy clients. In the original study, the fotonovela was provided to the caregiver with simple instructions to read the contents and attend one informational meeting to discuss contents in more detail. Caregivers were also followed up via phone calls to check whether they read the fotonovela, shared with their family or used suggested resources. Most Latino caregivers shared the booklet with other family members & attended the 1.5-hour group meeting to learn more about how the information could be personalized to their situation. ¡Unidos Podemos! has been shown to reduce depressive symptoms among Latino caregivers in a small RCT.^35^

Community health workers (CHWs) are ideal to deliver ¡Unidos Podemos! in primary care. Primary care is an optimal setting for identifying Latino caregivers of IWDs and delivering caregiver support programs as most older Latinos receive their care in primary care rather than specialty care. ^36,37^ Unfortunately, most primary care clinics offer little in terms of caregiver support for the caregivers of their patients with dementia due in part to providers’ lack of time and cultural proficiency.^38,39^ CHWs are being increasingly used to provide culturally tailored health education and connections with community resources in many primary care clinics.^40–42^ Thus, engaging CHWs to provide caregiver support programs among Latinos in primary care is a unique opportunity. In this manuscript we aim to identify barriers and facilitators to the implementation of ¡Unidos Podemos! by CHWs among Latinos in primary care. This research is crucial because there is a lack of robust evidence to inform the successful implementation of existing evidence-based ADRD caregiver support programs, especially among underserved communities.^43^ Studies to explore barriers and facilitators to the implementation of these programs could prove helpful in clinical practice.

## METHODS

### Design and participants

This study used a qualitative design and collected data via surveys and semi-structured interviews to identify barriers and facilitators to the implementation of ¡Unidos Podemos! by CHWs among Latinos in primary care. Clinic eligibility included primary care clinics in the US serving more than 100 Latinos 65 and older, with at least one bilingual community health worker or person with a similar educational and community-liaison role on staff. Participants were eligible if they worked in eligible clinics, were 18 or older, and worked as a CHW, a CHW supervisor, a primary care provider (PCP) or any other staff member the clinic deemed useful in implementing a caregiver program. We estimated a sample size of 20 participants to provide sufficient data to identify meaningful patterns and themes. The research team purposively recruited the clinics via their existing contacts and identified participants via clinic referral based on predetermined characteristics (at least one CHW and one PCP per clinic). The University of Kansas Medical Center Institutional Review Board approved this project (STUDY00149155). Before the survey, all participants completed an informed oral consent, which was written in the survey and read again before starting the interview. Clinics were offered $1,000 in compensation either as a payment or in-kind materials of equal value for the time of their workforce and individual interviewees were offered a $30 gift card.

### Procedures

Interviewers first contacted clinic leaders via email to schedule a follow-up videocall explaining the study’s background, purpose, compensation, and next steps. Interested clinics connected with their financial teams to arrange clinic compensation and referred eligible staff. Potential participants received an email outlining the study information. Eligible individuals were invited to complete an electronic survey on sociodemographic and clinic characteristics. After completing the survey, participants were contacted to schedule an interview. Those who completed the interview were considered enrolled in the study. Two study team members conducted all interviews between July and September of 2024 (JPP and MVRD). Interviews were conducted either in English or Spanish and recorded via a manual audio-recorder and the videocall platform recorder, to ensure full data capture in case one of the recorders failed. All interviews were transcribed and Spanish interviews were translated into English.

### Instruments

The instruments included one survey and one interview. The survey included basic demographic information, including gender, age, level of spoken Spanish and English (5-point Likert scale ranging from very low to very high), ethnicity and race. The survey also included clinic-related questions, including the location of the clinic (town, state), type of clinic (academic, safety net, federally qualified health clinic, private, other), and staff members’ role in the clinic (CHW, PCP, CHW supervisor, informatics staff, other healthcare staff, other administrative staff).

The interview guide, consisting of a 3-5 minute audio-visual description of the ¡Unidos Podemos! program and 8-10 open-ended questions, was developed based on the constructs the team considered key from all five domains of the Consolidated Framework for Implementation Research (CFIR) using the CFIR Interview Guide Tool (https://cfirguide.org/guide/app/#/).^44^ CFIR provides a prescriptive series of steps summarizing how implementation should ideally be planned and carried out. This framework includes five validated domains considered to be influential moderators or mediators of implementation outcomes.^45^ The domains include the: 1) individual characteristics, 2) characteristics of the program, 3) outer setting, 4) inner setting, and 5) implementation process.^44^ Appendix 1 contains the interview guides. To bring rigor and validity to the research process, the interviewers used active listening techniques during the interview aimed at confirming the information shared by the participants. They also emphasized the fact that participants were the experts in their experiences, to reduce power differentials. The team used detailed notes and memos to refine interview questions, interpret responses, and identify areas for further inquiry.^46^

### Data Analysis

Sample characteristics were summarized using SPSS v29.0.^47^ We conducted deductive qualitative analysis to analyze the interviews.^48^ First, we analyzed the data based on a priori constructs from the CFIR framework.^44^. We integrated the CFIR codebook schema (https://cfirguide.org/wp-content/uploads/2019/08/cfircodebooktemplate10-27-2014.docx)^44^ into Dedoose software, which includes all CFIR constructs organized by each of its five domains.^49^ Second, two coders systematically coded and interpret responses from 5% of respondents and compared code interpretation and application. Third, one coder systematically coded the remainder of the transcripts. Fourth, the second coder reviewed the remainder of the coding for agreement. Disagreements were reconciled by consensus.

## RESULTS

### Characteristics of the sample

Figure 1 shows the flow of clinics and participants identified and enrolled, as well as their reasons for exclusion. We initially identified 13 clinics and enrolled six. From these clinics, we enrolled 20 participants, in line with our recruitment goal. Table 1 shows the characteristics of the sample, which identified as largely women (n=15; 75.0%), and Latino (n=16; 80.0%).

**Figure 1:**
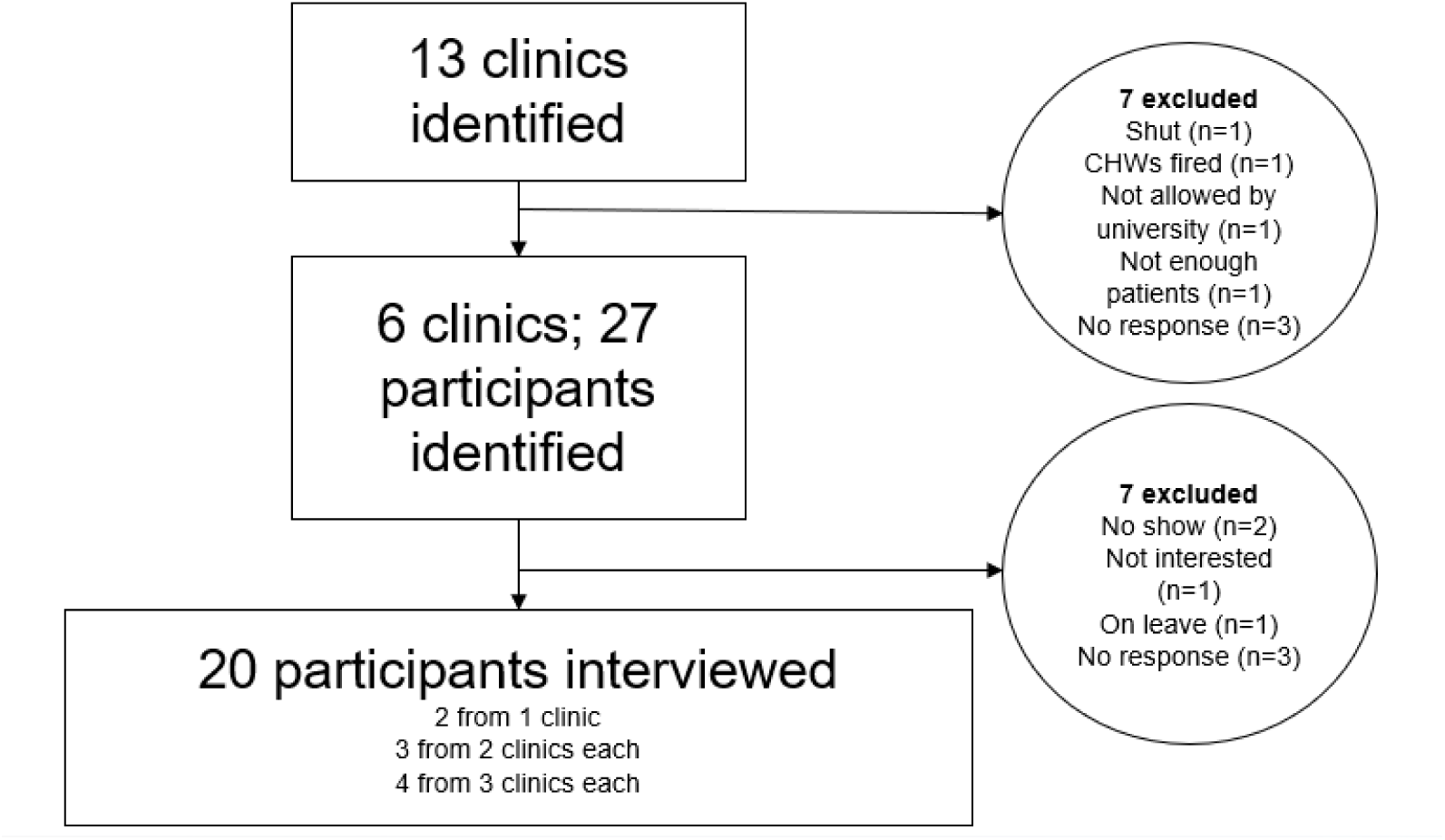
Participation flowchart

**Table 1.** Characteristics of participants.

|  | **% or $\mu$** | ***N* or**<br>***SD*** |
| --- | --- | --- |
| Women | 75.0% | 15 |
| Age | 41.3 | 9.8 |
| Ethnoracial group | 40.0% | 8 |
| White Latino | 35.0% | 7 |
| Other Latino | 5.0% | 1 |
| Mixed Latino | 10.0% | 2 |
| Non-Latino white | 5.0% | 1 |
| Non-Latino Black | 5.0% | 1 |
| Non-Latino Asian |  |  |
| Spoken Spanish (high or very high) | 60.0% | 12 |
| Spoken English (high or very high) | 95.0% | 19 |
| Rural practice | 30.0% | 6 |
| State | 35.0% | 7 |
| KS | 20.0% | 4 |
| PR | 20.0% | 4 |
| CA | 15.0% | 3 |
| MO | 10.0% | 2 |
| MI |  |  |
| Type of clinic |  |  |
| FQHC/Safety net | 45.0% | 9 |
| Private | 20.0% | 4 |
| Teaching health center | 20.0% | 4 |
| Academic | 15.0% | 3 |
| Role | 45.0% | 9 |
| CHW/supervisor | 30.0% | 6 |
| PCP | 15.0% | 3 |
| Other healthcare staff | 10.0% | 2 |
| Other admin staff |  |  |

Interviewees were diverse with respect to their spoken Spanish proficiency, rural/urban and state geography, and type of clinic they practiced in. Nine participants identified as a CHW or a supervisor (45.0%), six as a PCP (30.0%) and five as other staff (25.0%).

### Domain Themes

Within the five pre-established CFIR constructs described earlier, 21 themes were identified. Table 2 shows the themes and representative quotes of primary care clinic staff with respect to the implementation of ¡Unidos Podemos! in their clinics.

**Table 2:** Themes and representative excerpts of primary care clinic staff with respect to the implementation of ¡Unidos Podemos! in their clinics, following constructs from the five CFIR domains.

| **Individual Characteristics** |  |
| --- | --- |
| Beliefs and attitudes | *-“I would have loved to have this today.” PCP.*<br>*-“Caregivers are generally neglected. Nobody pays attention to them. It's always the patient. The caregiver must have the necessary education to be able to care for their family member. I think that it would be a feasible and useful program for my patients and their caregivers.” PCP.* |
| Self-efficacy | *-“If I see the fotonovela, I know that I could do it, because I've been working with this community already.”-CHW*<br>*-“We're small enough where we can identify patients via electronic health records. We've done a few other things like that”. PCP* |
| **Characteristics of the Program** |  |
| Relative advantage | *-“I feel like being specifically geared towards our patient population is what makes the difference between this and anything else they may see in the internet.” Clinic Coordinator*<br>*-“We don't have anything like this... Pictorial speaks better than just bunch of writeups in a paragraph.”-CHW* |
| Design and packaging | *-“I love the psychoeducation. I have a background in literature, so I love the fact that narrative is how we are educating and soothing and creating community and connection because they're not alone.” PCP*<br>*-“When the caregiver comes in, many of them find it difficult to process that their mother or close relative is already losing their memory. Like they can't accept it. It's difficult for them. It would be good if they could read that and see themselves identified” -CHW* |
| Adapting and reducing complexity | *-“I think the hour-and-a-half meeting, who would run it, where would we have it, that would be tough for us. I think giving out the fotonovelas and identifying patients wouldn't be hard. It would be the follow-up. ” Medical Director and PCP*<br>*-“I think the follow up meeting would definitely be nice. It's just really tricky.”-CHW Supervisor*<br>*-“They feel more comfortable if it's the same person.” Medical Assistant* |
| **Outer setting** |  |
| External policy and rewards | *-“Remember that we are in a capitalist society. There has to be an incentive”-PCP.*<br>*-“Having services like this is valued in this setting. I just don't know how it'd be paid for.”-PCP* |
| Cosmopolitanism | *-“Where we live there is a very large farming population. They listen to the radio. This could be a way to reach them”-PCP.*<br>*-“We have a partner diaper program. We have food boxes available also here on hand.”-CHW* |
| Patient needs and resources | *-“They have different schedules. There is a lot of variability among field workers, depending on what type of crop they have (carrots, grapes, pistachios). Work in the summer is a little more complicated than in winter* |
|  | *because in winter they have more time available.”-PCP*<br>*–“The cultural and social stigma is always very strong. Perhaps they are a little reluctant and they refrain from seeking help. Because of the stigma that has existed for a long time... about depression and about mental illnesses in general”-Clinic Coordinator* |
| **Inner Setting** |  |
| Available resources | *–“We have a team of four clinic-based community health workers that are all bilingual in English and Spanish. I also work with our behavioral health team. And currently we have four clinicians and three of those clinicians are bilingual in English and Spanish.” -CHW Supervisor*<br>*–“a barrier would be being able to have the staff and the time to implement any program into our clinic.”-Healthcare staff* |
| Access to knowledge and information | *–“If you can draft up a workflow, step by step, and maybe a script, so we can explain them to them ‘This is what it is. This is the benefits. This is what we're willing to offer you...’ Something visual would be best.”-Medical Assistant*<br>*–‘I want the community health worker to understand what dementia is, how it works, to be able to work with our client... That training that will show me how to look for other resources, to do the assessments, reports, and specifics of documenting.”-CHW* |
| Compatibility | *–“We're a residency program... they leave every three years. Somebody would have to pick it up.... to continue the program and make sure that it's thriving.” PCP*<br>*–“I think it will be beneficial because when we go we don't have anything like this”- CHW* |
| Culture | *–“We're always looking to better our patient population and their health. There's always a lot of stuff that can be implemented. It's always what is best for our patient population and can we handle it as a clinic.” -CHW*<br>*–“That's why I love working here. We all are a group and we're always looking for the best, or we're always looking out for the patient just to make sure that they're helped in every single aspect of their life.”-CHW* |
| Goals and feedback | *–“It usually has to have some ability to either show how many we’ve given out or, if there’s a positive impact with our patients”-PCP*<br>*–“ I would like a message back about what happened. Or at least be able to look in the chart and say, ‘Oh. Oh yeah. They met with this team, and these were the changes that occurred or this was the new program that they were enrolled in.’” PCP* |
| Networks and communication | *–“We have a meeting every morning that we call a huddle meeting. Announcements are made and these new initiatives are followed up through meetings that we have either on Wednesday afternoons or through huddles. That's the way we implement new things.”-PCP*<br>*–“We receive the referrals from the different clinics over our electronic system that we use. And some we get via email, some we get via a Teams message. -CHW* |
| Tension for change | *–“We have been seeing a lot more patients with dementia.”-Medical Assistant* |
|  | -“I want to call the patient I met today, this husband whose wife has significant memory loss and sundowning. This is what they need.”-PCP |
| Incentives and rewards | -“That's my job. So our incentive is my clients get their goals.”-CHW<br>-“I don't think incentives are necessary. It would be part of your mission and passion. The reason why you come to work every day is to help our patients live a healthier, sustainable life. If the program was implemented, it would just go hand in hand.” CHW |
| **Implementation Process** |  |
| Champions | -“It could be the MDs. But the health personnel within the clinic can also do it, although I really feel that we as doctors would be a little more appropriate because we could also talk to them a little more fully about what it is and what we want to achieve with them” – Coordinator<br>-“Everything would have to be ran by our Chief Medical Officer.”-CHW |
| Engagement | -“I think it would just be multiple meetings with just really good information and something that we are able to bring back that we can say we're ready to go. In between both in-person and through Zoom.”-CHW<br>-“If you can draft up a workflow or something, just step by step of how to – maybe a script and then utilizing the resources so we can explain them to them; ‘This is what it is. This is the benefits. This is what we're willing to offer you’”-CHW |
| Execution | -“In my mind, it could be a community health worker who calls and says ‘What are you thinking about?’, ‘Have you looked at the book again? ‘Have you guys thought about daycare or self-care?’.” PCP<br>-“I think it would be helpful that the community health worker presented it to them. They review the fotonovela. Not page by page, but this first part covers blah, blah, blah. And then this covers blah, blah, blah. Giving an overview while flipping through it.”-CHW supervisor |
| Formally appointed internal implementation leaders | -“I think that would be the community health workers (to identify participants). I think we as the workers do try to communicate everything to our patients.”-CHW<br>-“This person would have to be a medical assistant who is dedicated to that. A medical assistant. The person making this call”-PCP |
| Reflecting and evaluating | -“When we identify someone that needs a resource we need to know if it will be the correct person for that program. I don't like my clients to be rejected, so make sure that this service is working, and it will work for them.” CHW<br>-“We need to make sure that they're actually being taken care of and actually using the resources that are given to them. And not just pushed to the side because I know a lot of people that are given a pamphlet and they just take it, go home and it just stands on the table. But if the phone calls are being done and you're checking in, at least you're making sure that the patient is actually doing it.” -Administrative staff |

#### Individual Characteristics

Participants had positive <u>beliefs and attitudes</u> toward the proposed program, desired more time with the program to familiarize themselves, and felt that this program could be implemented in their setting. All participants saw the value of this program because, to them, the dearth of attention for Alzheimer’s patients’ caregivers was unproductive, unhelpful, and neglectful. Regarding <u>self-efficacy</u>, many of the participants felt very capable of executing an implementation if given the right amount of education, training, materials, and tools.

#### Characteristics of the Program

Participants stated that this program has a <u>relative advantage</u> over standard care, as there was nothing available at their settings at present to meet the needs of Spanish speaking caregivers of dementia patients. Regarding the <u>design and packaging</u>, participants liked the comic-like packaging of the fotonovela, its pictoriality and simplicity, the fact that it begins with a description of dementia, and the focus on Latinos. Participants stressed the importance of <u>adapting the</u> <u>delivery and reducing complexity</u> of the components that supplement the fotonovela: 1.5 hour in-person group meeting, and multiple follow-up calls. Adaptations should aim at reducing the time and transportation needs of caregivers and staff. These adaptations could include reducing the group discussion meeting’s duration and the frequency of follow-up calls and being flexible in allowing in-person vs remote modalities. Adapting the delivery also included the same staff member being involved in all components. This involvement would provide a sense of continuity that is important to increase adherence due to trust and fewer risk of caregiver falling through the cracks of administrative errors.

#### Outer setting

A theme within the outer setting was <u>external policy and rewards</u>. Despite the high implementability of delivering the fotonovela to caregivers, participants spoke of the importance of the components that supplement the fotonovela (group discussion meetings and follow-up calls) to be covered by insurance or some sort of method of financial support. Participants discussed their existing networks with external organizations (<u>cosmopolitanism</u>) to meet caregivers and clinic needs. These networks included social workers or the Alzheimer’s association to deliver components of the program (e.g., group meeting), agencies to deal with social determinants of health (transportation, food insecurity, housing insecurity, utility assistance, or translation), or media groups to promote services. Participants suggested that the following <u>patient needs and resources</u> should be considered for the implementation of the program: memory loss care, Spanish language materials for people with low literacy and culturally/language-competent staff, adapting to caregivers’ complex work schedules, and addressing stigma, which is common among Latinos and can impact sharing their emotions with a group.

#### Inner Setting

<u>Available resources</u> included Spanish-speaking staff, and electronic health records that could be leveraged to identify caregivers via their loved ones with dementia and keep track of their participation in the program. All participants stated that, to their knowledge, there is no current tool for caregiver support. Participants highlighted their desire for a step-by-step decision support tool with graphic and generic descriptions that could be expanded into a more detailed document as needed, to increase <u>access to knowledge and information</u>. Regarding <u>compatibility</u>, participants stated agreement with the values of the program meeting the needs of underserved caregivers of dementia. However, given the turnover of staff in some clinics, some worried about the program’s fit into clinic workflows. Regarding <u>culture</u>, all participants spoke of their clinics as valuing the humanity and dignity of their patients, striving to meet their needs, and serving as a respectful member of the community. Participants spoke of the importance of gaining their patient’s trust and good stewardship of that trust once they had it. Participants spoke of the need for <u>goals and feedback</u> on delivery and outcomes to sustain the program and keep sight of measurable goals, as well as show granting organizations the work done in the clinic.

Most participants described a potential process of <u>networks and communication</u> about patient needs, dementia and other health statuses, supports, and protocols using electronic health records. Participants reported identifying undiagnosed patients and their caregivers via informal communication such as waiting for the patient or caregiver to raise a concern about their cognition, while others had formal screening processes. All participants described recurrent meetings where feedback and information about the clinic and its goals were shared. Most participants saw the current lack of support for caregivers as untenable and in direct need for social support, creating a <u>tension for change</u>. Most statements about <u>incentives and rewards</u> for staff were that coming to work, doing their job, and helping patients were their incentives.

#### Implementation Process

Most participants saw the PCPs as the potential <u>champions</u>, as they would have the most sway regarding the continued support of patients, caregivers, and other clinic staff, and have a high degree of authority. However, some also mentioned that the Chief Medical Officer needed to be on board for program implementation. Regarding <u>engaging</u>, participants stated that with enough training, implementation of the fotonovela program would be successful. They suggested multiple methods of training, including in-person or remote modalities, brochures, videos, and decision guides. They iterated the importance of either horizontal or vertical supervision and recurrent reminders to be a key factor in predicted success of implementation. Most participants thought the <u>execution</u> of most components of the program should be conducted by CHWs. CHWs were seen as the most likely to have the time, resources, and knowledge of the patients and their caregiver’s situation. For this reason, they were also <u>formally appointed internal implementation leaders</u>. Medical assistants were seen as an alternative to CHWs. Regarding <u>reflecting and evaluating</u>, participants would like regular check-ins with their team about the fit of the implementation in their clinic, and checks and balances so that caregivers do not fall through the cracks administratively or get involved in a program that is not appropriate. Participants also spoke of wanting to collect some data on how their patients benefit from the program.

## DISCUSSION

This study aimed to identify barriers and facilitators to the primary care implementation of an evidence-based dementia caregiver support programs tailored to Latinos by CHWs. Findings showed the potential value of ¡Unidos Podemos!, given the simple implementability of delivering the fotonovela, its culturally-appropriate design and packaging, and the fact that, despite there being evidence-based programs for the general and the Latino population,^50^ these are not being implemented in primary care.^51^ Factors that could increase the implementability of the program refer to the components that are supplemental to the fotonovela, and include reimbursement and the time and transportation caregiver and staff needs to deliver these supplemental components, and the need for training, decision support tools, supervision and feedback mechanisms.

All participants had positive attitudes towards ¡Unidos Podemos!, especially the tailored format of the fotonovela and the content. In fact, participants considered this program unique, given that their clinic were providing no alternatives for Latinos. These findings are consistent with previous literature showing that primary care clinics rarely serve caregivers directly and have access to caregiver support tools.^51^ These reactions are consistent with the current scarcity of implementation science trials on ADRD caregiver support programs.^43,52^ These findings also suggest that the existing evidence-based interventions for Latinos, which are very diverse with respect to delivery modality,^50^ should be further tailored to primary care clinic settings, and promoted better.

Participants thought the execution of most components of the program should be conducted by CHWs, as they have the time, resources, and knowledge of the patients and their caregiver’s situation. Moreover, currently at thirty percent, a growing number of community health clinics, which disproportionately serves Latinos, hire CHWs.^42^ Despite this, our findings suggest that other staff can also be good candidates for delivering this program, including medical assistants. Primary care clinics will require PCPs in most cases to champion the implementation. The importance of PCPs to champion dementia care is in line with a growing trend of work acknowledging that PCPs are the most ideal gatekeepers for patients with dementia, which can open the door to screening, diagnosis, treatment, care, and research.^37^

Despite the perceived high implementability of delivering the fotonovela, findings suggest opportunities to increase the implementation and sustainability of ¡Unidos Podemos! These opportunities refer to supplemental components: the 1.5 hour in-person group meeting, and follow-up calls. One of these opportunities is identifying sources of reimbursement for these components. For example, if supplemental components are to be implemented by CHWs, clinics can bill services using the 2024 Community Health Integration under the Centers for Medicare and Medicaid Services (CMS).^53^ These codes include in-person or virtual emotional support and navigation services provided by a certified CHW under the direction of a practitioner. Another opportunity is shortening the group discussion’s duration, reducing the frequency of phone calls, and leveraging different technologies to reduce time and transportation needs.^54–56^ New programs have leveraged technologies such as videocalls and text messaging that could be integrated to increase the feasibility of delivery of ¡Unidos Podemos! components.^57–62^ Together, these opportunities raise the question about whether these components are core or peripheral, this is, whether providing the fotonovela with a brief overview and suggestion to read it and share it with the family would be effective, or whether the supplemental group discussion and follow-up calls are necessary. In the original study where ¡Unidos Podemos! was found to be efficacious, most participants in both the fotonovela and the control group attended group discussion meetings.

This study has limitations, which are linked to the generalizability of our findings. First, most interviewees identified as Latino. While Latino PCPs might be more likely to serve Latino patients, many Latino patients receive care from PCPs with other backgrounds. Second, this study excluded clinics with 100 or fewer older Latino patients, and those without at least one bilingual CHW. The sample size was relatively small and non-probabilistic. The capabilities and needs of clinics that were not included for these reasons might differ from the ones included in these findings.

Findings from this study have direct implications for practice and public health. Many primary care clinics are equipped and see a need to provide caregiver support to Latinos. However, these would need the tool to do so, and ¡Unidos Podemos! might be a unique opportunity, especially if implementation strategies address the determinants found in this manuscript. This program could be integrated within state-wide CHW training events in states that serve large Latino communities. If supplemental components are to be integrated in addition to providing the fotonovela, implementation scientists and practitioners need to start discussions on how this implementation should be funded, especially in clinics that serve uninsured Latino caregivers. Future studies should disentangle the separate effects of providing the fotonovela vs supplementing it with components that might be harder to implement such as group discussion meetings and follow-up calls using for example, Multiphase Optimization STrategy (MOST) designs.^63^ Future studies should develop and test implementation strategies to deliver ¡Unidos Podemos! in primary care. A feasible path would first be pilot testing a strategy, and only if this strategy and the design are feasible, conduct a fully-powered trial to test the effectiveness and implementation outcomes, given that ¡Unidos Podemos! has not yet been tested for effectiveness. Studies could aim to assess barriers and facilitators of implementing ¡Unidos Podemos! in other settings that are likely to identify Latinos with memory disorders, including churches. More studies should aim to assess barriers and facilitators to other evidence-based caregiver support programs for Latinos. Especially those with remote and asynchronous components, which have large potential to reach large amounts of Latinos.^50^

## CONCLUSION

This study has identified barriers and facilitators to implementing an evidence-based dementia caregiver support program among Latinos. ¡Unidos Podemos! is seen as a unique and mostly-simple to implement program. To further improve the implementability of this program, implementers might want to address the components that supplement the fotonovela, namely the group discussion meeting and follow-up calls. Addressing these components include identifying sources of reimbursement for these services, and developing and providing staff training, decision support tools, supervision and feedback. Future research could help clarify which of the program components is crucial vs peripheral and develop implementation strategies that address these determinants. Addressing these determinants would constitute one of the first ever approaches to test the implementation of a caregiver support strategy among Latinos and bring the evidence about eliminating Latino caregivers’ disproportionate impact to the real world.

## Data Availability

All data produced in the present study are available upon reasonable request to the authors.

## Appendix 1. Interview guides

### Only for Community Health Workers and their managers

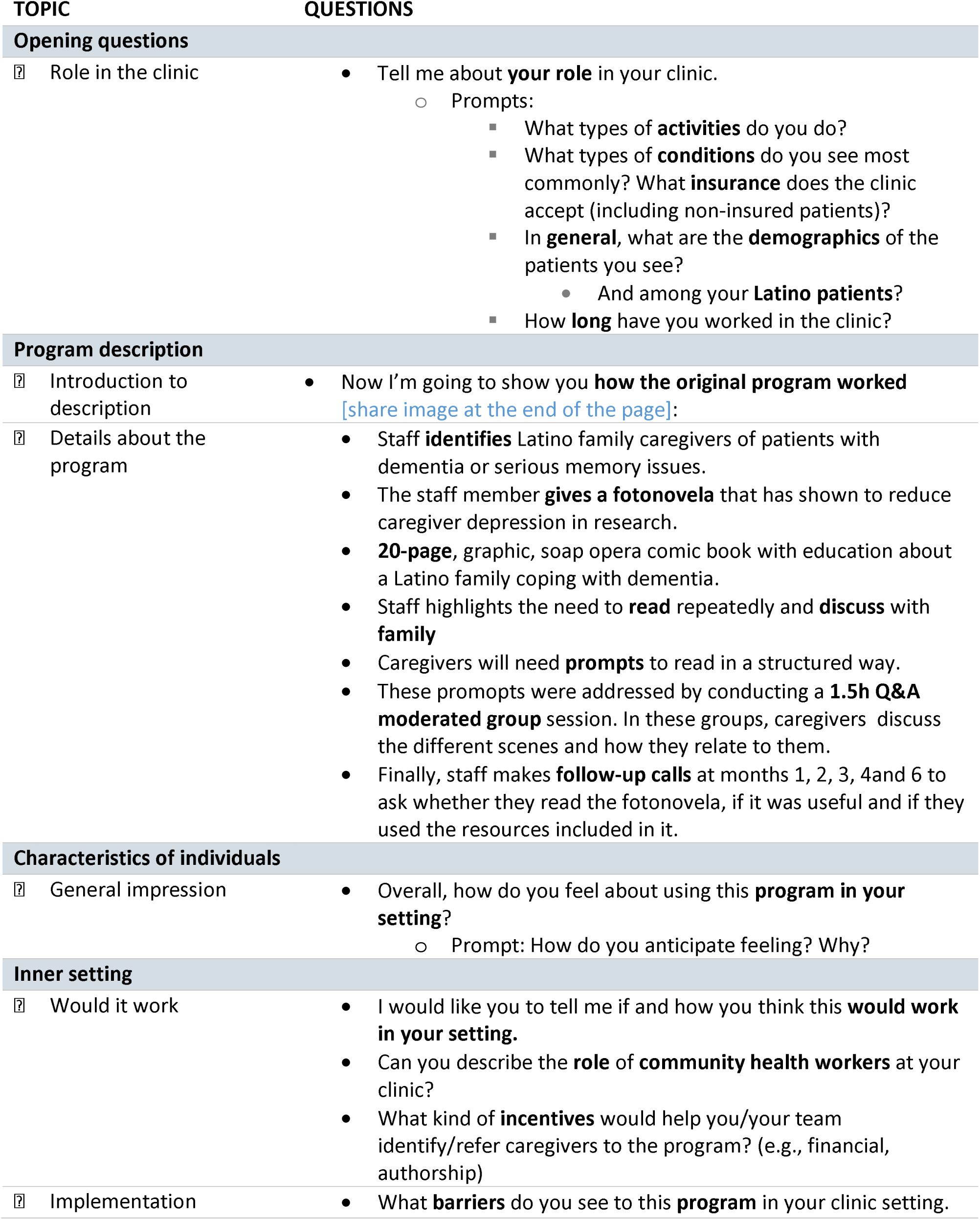
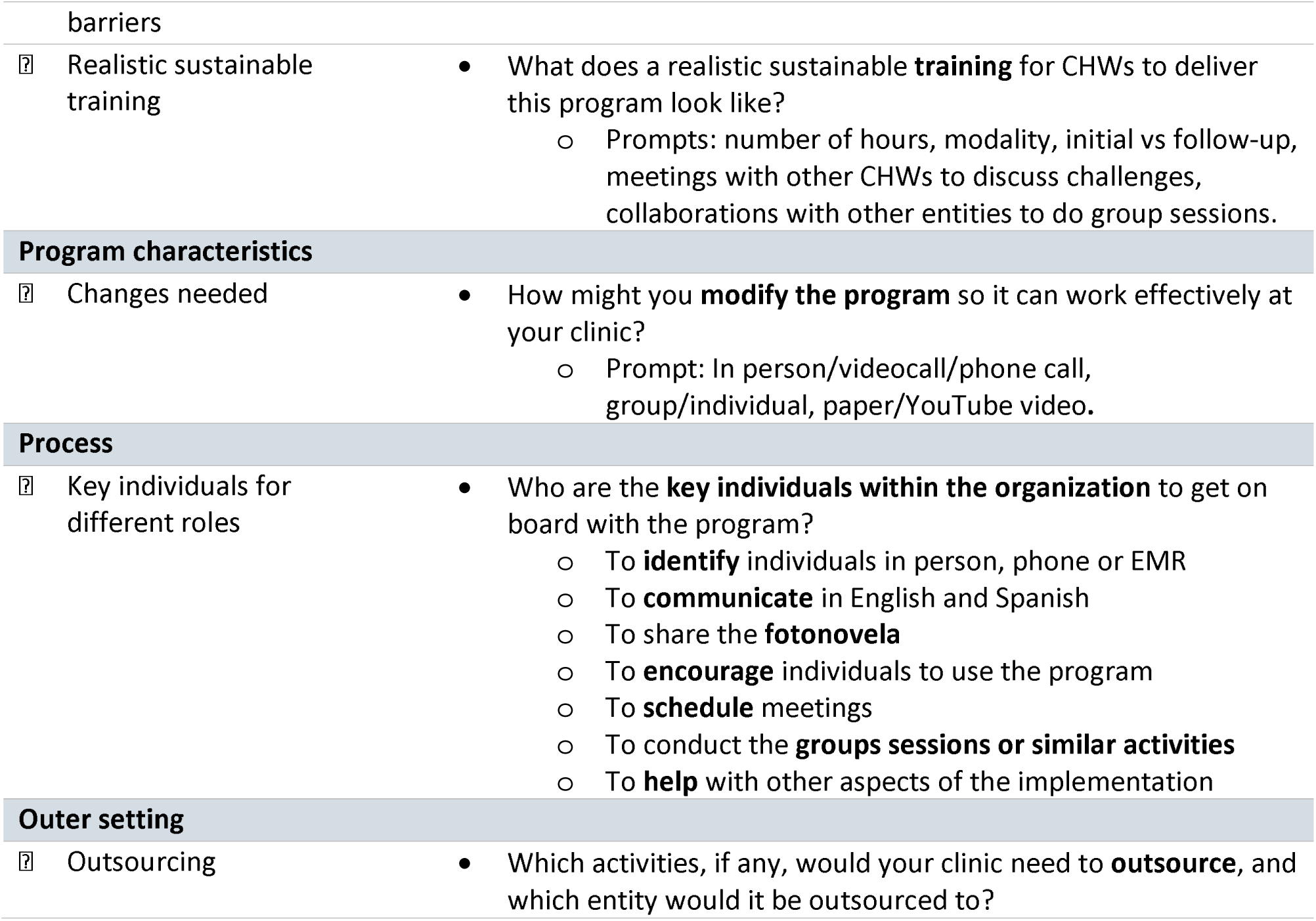

Appendix 1. Interview guides. Only for Community Health Workers and their managers
| TOPIC | QUESTIONS |
| --- | --- |
| **Opening questions** |  |
| ☐ Role in the clinic | • Tell me about **your role** in your clinic.<br>○ Prompts:<br>▪ What types of **activities** do you do?<br>▪ What types of **conditions** do you see most commonly? What **insurance** does the clinic accept (including non-insured patients)?<br>▪ In **general**, what are the **demographics** of the patients you see?<br>• And among your **Latino patients**?<br>▪ How **long** have you worked in the clinic? |
| **Program description** |  |
| ☐ Introduction to description | • Now I'm going to show you **how the original program worked** [share image at the end of the page]: |
| ☐ Details about the program | • Staff **identifies** Latino family caregivers of patients with dementia or serious memory issues.<br>• The staff member **gives a fotonovela** that has shown to reduce caregiver depression in research.<br>• **20-page**, graphic, soap opera comic book with education about a Latino family coping with dementia.<br>• Staff highlights the need to **read** repeatedly and **discuss** with **family**<br>• Caregivers will need **prompts** to read in a structured way.<br>• These prompts were addressed by conducting a **1.5h Q&A moderated group** session. In these groups, caregivers discuss the different scenes and how they relate to them.<br>• Finally, staff makes **follow-up calls** at months 1, 2, 3, 4 and 6 to ask whether they read the fotonovela, if it was useful and if they used the resources included in it. |
| **Characteristics of individuals** |  |
| ☐ General impression | • Overall, how do you feel about using this **program in your setting**?<br>○ Prompt: How do you anticipate feeling? Why? |
| **Inner setting** |  |
| ☐ Would it work | • I would like you to tell me if and how you think this **would work in your setting**.<br>• Can you describe the **role** of **community health workers** at your clinic?<br>• What kind of **incentives** would help you/your team identify/refer caregivers to the program? (e.g., financial, authorship) |
| ☐ Implementation | • What **barriers** do you see to this **program** in your clinic setting. |

|  |  |  |
| --- | --- | --- |
| barriers |  |  |
| ? | Realistic sustainable training | What does a realistic sustainable **training** for CHWs to deliver this program look like?<br>Prompts: number of hours, modality, initial vs follow-up, meetings with other CHWs to discuss challenges, collaborations with other entities to do group sessions. |
| **Program characteristics** |  |  |
| ? | Changes needed | How might you **modify the program** so it can work effectively at your clinic?<br>Prompt: In person/videocall/phone call, group/individual, paper/YouTube video. |
| **Process** |  |  |
| ? | Key individuals for different roles | Who are the **key individuals within the organization** to get on board with the program?<br>To **identify** individuals in person, phone or EMR<br>To **communicate** in English and Spanish<br>To share the **fotonovela**<br>To **encourage** individuals to use the program<br>To **schedule** meetings<br>To conduct the **groups sessions or similar activities**<br>To **help** with other aspects of the implementation |
| **Outer setting** |  |  |
| ? | Outsourcing | Which activities, if any, would your clinic need to **outsource**, and which entity would it be outsourced to? |

### Only for Administrative staff and PCPs

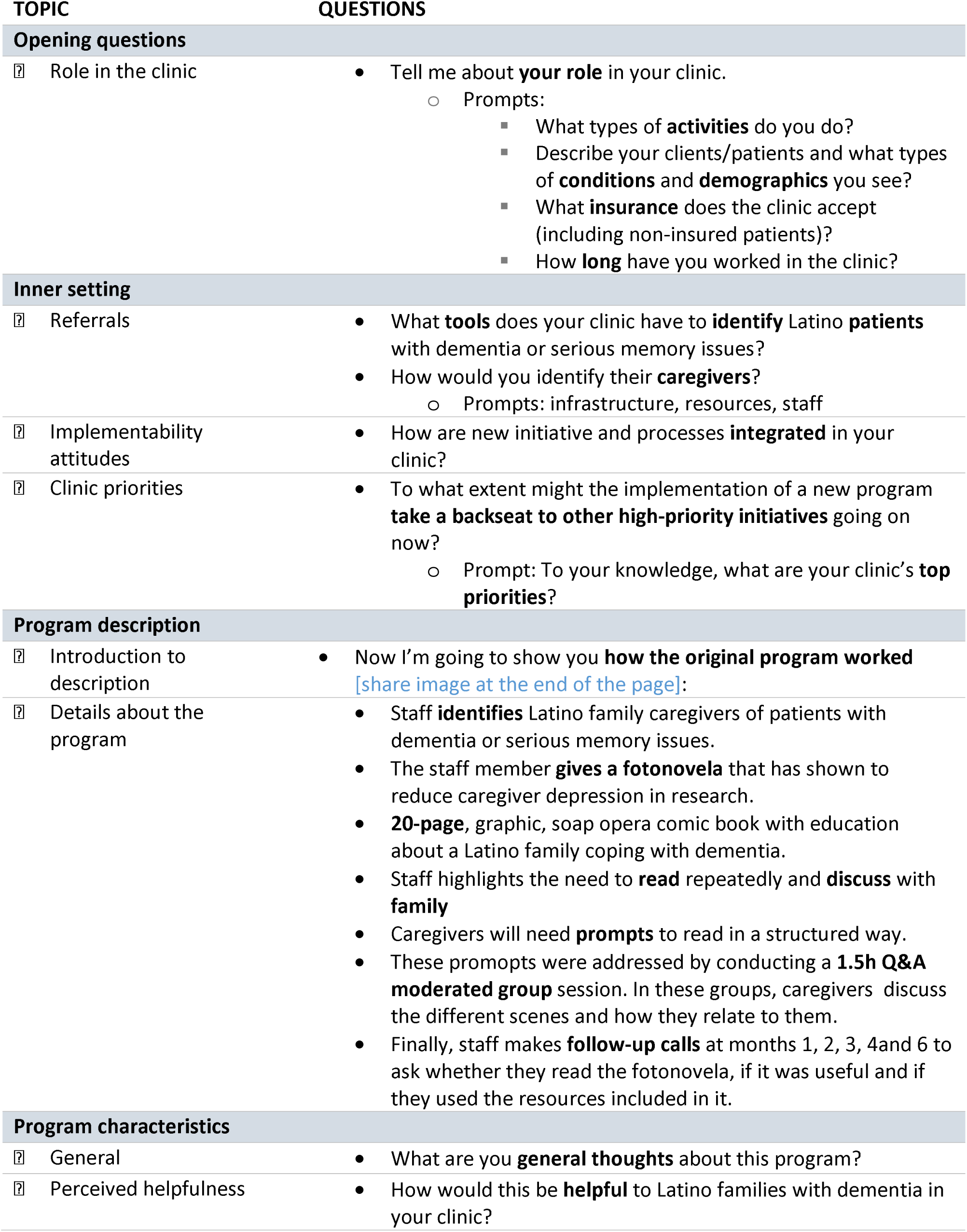
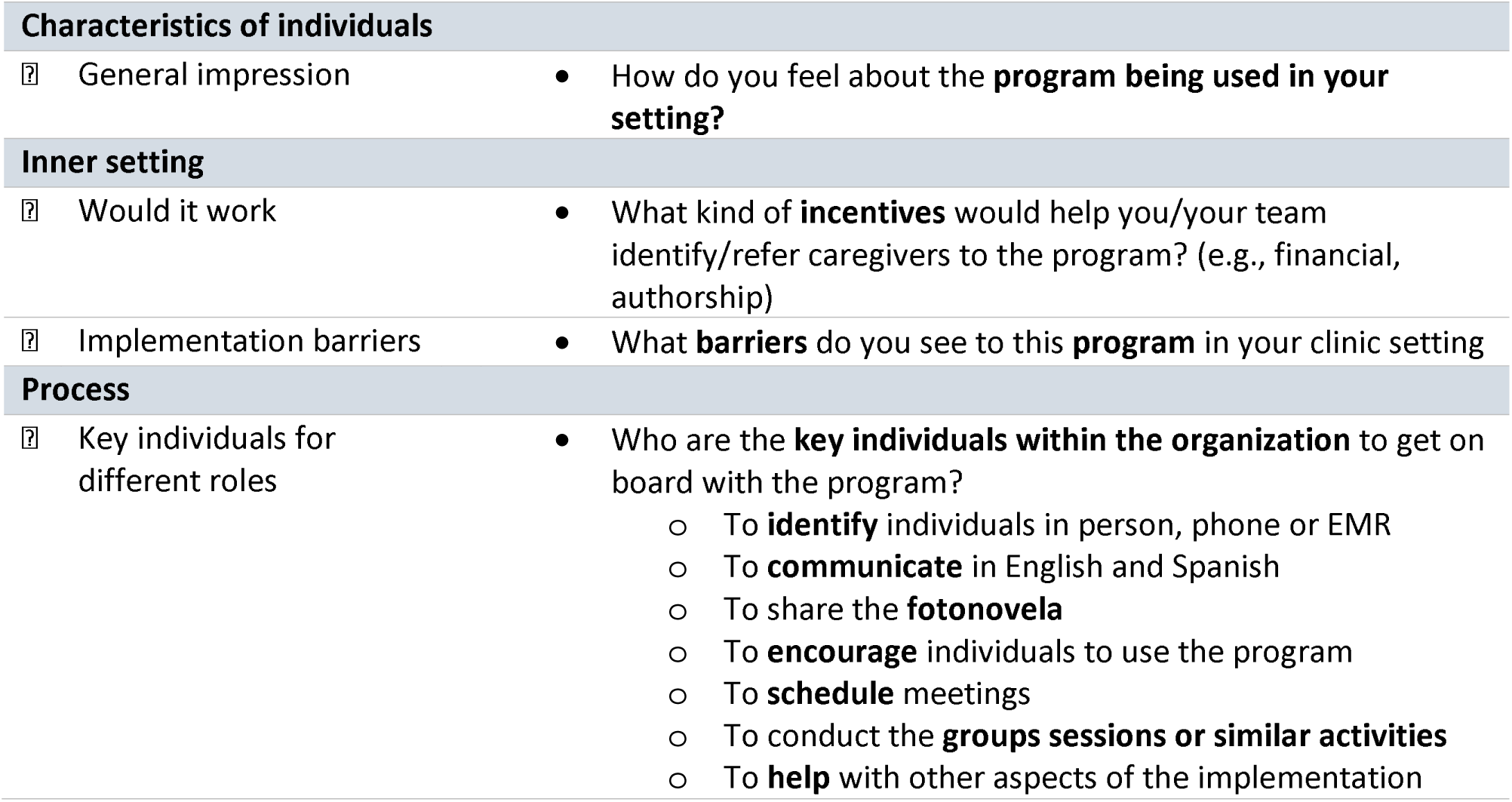

| TOPIC | QUESTIONS |
| --- | --- |
| **Opening questions** |  |
| ☐ Role in the clinic | Tell me about **your role** in your clinic.<br>Prompts:<br>What types of **activities** do you do?<br>Describe your clients/patients and what types of **conditions** and **demographics** you see?<br>What **insurance** does the clinic accept (including non-insured patients)?<br>How **long** have you worked in the clinic? |
| **Inner setting** |  |
| ☐ Referrals | What **tools** does your clinic have to **identify** Latino **patients** with dementia or serious memory issues?<br>How would you identify their **caregivers**?<br>Prompts: infrastructure, resources, staff |
| ☐ Implementability attitudes | How are new initiative and processes **integrated** in your clinic? |
| ☐ Clinic priorities | To what extent might the implementation of a new program **take a backseat to other high-priority initiatives** going on now?<br>Prompt: To your knowledge, what are your clinic's **top priorities**? |
| **Program description** |  |
| ☐ Introduction to description | Now I'm going to show you **how the original program worked** [share image at the end of the page]: |
| ☐ Details about the program | Staff **identifies** Latino family caregivers of patients with dementia or serious memory issues.<br>The staff member **gives a fotonovela** that has shown to reduce caregiver depression in research.<br>**20-page**, graphic, soap opera comic book with education about a Latino family coping with dementia.<br>Staff highlights the need to **read** repeatedly and **discuss** with **family**<br>Caregivers will need **prompts** to read in a structured way.<br>These prompts were addressed by conducting a **1.5h Q&A moderated group** session. In these groups, caregivers discuss the different scenes and how they relate to them.<br>Finally, staff makes **follow-up calls** at months 1, 2, 3, 4 and 6 to ask whether they read the fotonovela, if it was useful and if they used the resources included in it. |
| **Program characteristics** |  |
| ☐ General | What are you **general thoughts** about this program? |
| ☐ Perceived helpfulness | How would this be **helpful** to Latino families with dementia in your clinic? |
| ☐ General impression | • How do you feel about the **program being used in your setting?** |
| ☐ Would it work | • What kind of **incentives** would help you/your team identify/refer caregivers to the program? (e.g., financial, authorship) |
| ☐ Implementation barriers | • What **barriers** do you see to this **program** in your clinic setting |
| ☐ Key individuals for different roles | • Who are the **key individuals within the organization** to get on board with the program?<br>○ To **identify** individuals in person, phone or EMR<br>○ To **communicate** in English and Spanish<br>○ To share the **fotonovela**<br>○ To **encourage** individuals to use the program<br>○ To **schedule** meetings<br>○ To conduct the **groups sessions or similar activities**<br>○ To **help** with other aspects of the implementation |

